# Association between polygenic propensity for a psychiatric disorder and nutrient intake

**DOI:** 10.1101/2021.01.18.21249750

**Authors:** Avina K. Hunjan, Christopher Hübel, Yuhao Lin, Thalia C. Eley, Gerome Breen

**Affiliations:** Social Genetic & Developmental Psychiatry Centre, Institute of Psychiatry, Psychology & Neuroscience, King’s College London, UK; NIHR Biomedical Research Centre for Mental Health; South London and Maudsley NHS Trust, UK; Department of Medical Epidemiology and Biostatistics, Karolinska Institutet, Stockholm, Sweden; National Centre for Register-based Research, Department of Economics and Business Economics, Aarhus University, Aarhus, Denmark

## Abstract

**Background:** Despite the observed associations between psychiatric disorders and nutrient intake, genetic studies are limited.

**Aims:** We examined whether polygenic scores for psychiatric disorders, including anorexia nervosa, major depressive disorder and schizophrenia, are associated with self-reported nutrient intake.

**Methods:** We used data obtained by the UK Biobank ‘Diet by 24-hour recall’ questionnaire (N=163,619). Association was assessed using linear mixed models for the analysis of data with repeated measures.

**Results:** We find polygenic scores for psychiatric disorders are differentially associated with nutrient intake, with attention-deficit/hyperactivity disorder, bipolar disorder and schizophrenia showing the strongest associations, whilst autism spectrum disorder showed no association. Expressed as the effect of a one standard deviation higher polygenic score, anorexia nervosa polygenic score was associated with higher intake of fibre (0.06 g), folate (0.93 μg), iron (0.03 mg) and vitamin C (0.92 μg). Similarly, a higher major depressive disorder polygenic score was associated with 0.04 mg lower iron and 1.13 μg lower vitamin C intake per day, and a greater obsessive-compulsive disorder polygenic score with 0.06 g higher fibre intake. These associations were predominantly driven by socioeconomic status and educational attainment. However, a higher alcohol dependence polygenic score was associated with higher alcohol intake and individuals with higher persistent thinness polygenic scores reported their food to weigh 8.61 g less, both independent of socioeconomic status.

**Conclusions:** Our findings suggest that polygenic propensity for a psychiatric disorder is associated with dietary behaviour. The nutrient intake is based on self-reported data and findings must therefore be interpreted mindfully.

**Declaration of interest:** None.

## Introduction

Psychiatric disorders such as major depressive disorder (MDD), anxiety disorders, schizophrenia and attention deficit hyperactivity disorder (ADHD) affect 20-25% of the population at any one time (Gustavson et al. 2018; Charlson et al. 2019). These disorders are often accompanied by disturbances in food intake and nutritional status. For example, reduced protein intake has been associated with reduced cognitive functioning amongst individuals with schizophrenia (Dickerson et al. 2020). In addition, vitamin deficiencies is associated with psychiatric disorders: lower concentrations of vitamins B6 and B9 (i.e., folate) with an ADHD diagnosis, B6 with symptom severity of inattention, hyperactivity and impulsivity (Landaas et al. 2016); deficiencies in vitamins B9 and B12 with worse negative symptoms in schizophrenia (El Mawella, Hussein, and Ahmed 2018); and vitamin B12 deficiency with the development of obsessive-compulsive disorder (OCD) (Valizadeh and Valizadeh 2011; Esnafoğlu and Yaman 2017). Dietary deficiencies in iron, vitamin C and vitamin D are also common in individuals with a psychiatric diagnosis (Hoffer 2008; Bajpai et al. 2014; Gariballa 2014; Esnafoğlu and Yaman 2017; Bener and Kamal 2013; Belzeaux et al. 2015; Milaneschi et al. 2014; Chen et al. 2013). However, these observations merely demonstrate associations and to establish cause and effect relationships between the development and/or severity of psychiatric disorders and alterations in dietary behavior, more in-depth investigation of their co-occurrence is needed.

Both psychiatric disorders and nutrient intake are heritable. Psychiatric disorders are complex traits influenced by thousands of genetic variants; and genome-wide association studies (GWASs) have identified more than 300 independent genomic loci (Polderman et al. 2015; Schizophrenia Working Group of the Psychiatric Genomics Consortium 2014; Howard et al. 2019; Cross-Disorder Group of the Psychiatric Genomics Consortium 2019). Twin analyses have also revealed a high heritability for total energy intake (twin-*h*^2^ = 48%), macronutrients (35-45%; i.e. protein, carbohydrates and fats), minerals (45%; includes calcium, iron and potassium) and vitamins (21%; including vitamins A, C, D, E, and carotenoids, such as alpha and beta carotenes) (J. Liu et al. 2013). Six GWASs of dietary intake have been performed (Merino et al. 2018; Tanaka et al. 2013; Chu et al. 2013; Meddens et al. 2020; Reed et al. 1997; Cole, Florez, and Hirschhorn 2020), with the largest identifying 309 associated loci, indicating olfactory receptor genes associated with fruit and tea intake (Cole, Florez, and Hirschhorn 2020).

Despite i) the phenotypic evidence for an association between psychiatric disorders and disturbances in nutrient intake and ii) the heritability of both, little empirical attention has been given to understanding their genetic overlap. A recent UK Biobank study found significant genetic correlations between schizophrenia and two diet groups - one representing a meat-related diet and the other a fish and plant-related diet (Niarchou et al. 2020). This study highlights the need for more genetic studies to better understand the relationship between psychiatric disorders and nutrient intake. This is clinically important because unhealthy dietary habits can impact physical and psycho-social health (Polivy and Herman 2005) imposing further burden on individuals with a psychiatric disorder. Genetic studies could help determine whether development of integrative treatment strategies that target behaviour alterations in diet are needed to improve the long-term management of these disorders.

In this study, We explore the association between polygenic scores for psychiatric disorders and self-reported nutrient intake using data from the UK Biobank to determine whether polygenic propensity for a psychiatric disorder is associated with dietary behaviour. Specifically, we looked at polygenic scores for eight psychiatric disorders, including anorexia nervosa, schizophrenia, and major depressive disorder (MDD), as well as food addiction, persistent thinness, educational attainment, BMI, height, using systemic lupus erythematosus (lupus) as a negative control. We examined their association with the intake of fourteen nutrients derived from the UK Biobank ‘Diet by 24-hour recall’ questionnaire.

## Methods

### Ethics

We obtained approval for this research under an approved access request (application 23395) to UK Biobank. UK Biobank has approval from the North West Multi-centre Research Ethics Committee (MREC), which covers the UK, and the Patient Information Advisory Group (PIAG) for gaining access to information that would allow it to invite people to participate. Our use of the data was governed by the analysis plan in our access request and the terms of the material transfer agreement between KCL and UK Biobank. We assert that all procedures contributing to this work comply with the ethical standards of the relevant national and institutional committees on human experimentation and with the Helsinki Declaration of 1975, as revised in 2008.

### Study population, genotype quality control and sample size

The UK Biobank is a large prospective cohort study consisting of approximately 500,000 participants aged 46-69 years when recruited in 2006–2010 (Allen et al. 2014). Written informed consent was obtained from all participants. The study assessed dietary behaviour using a web-based 24-hour dietary assessment tool which asked about the frequency of consumption of common foods and drinks (Category 100090) (B. Liu et al. 2011) Participants were asked whether what they ate and drank yesterday was typical (Data-Field 100020) and if they routinely followed a special diet (Data-Field 20086). Responses were automatically coded to provide estimated daily nutrient intake (Category 100098). Participants completed the initial assessment at recruitment centres and then remotely on four occasions between April 2009 and June 2012 (see Supplementary Materials for further information).

Genome-wide genetic data came from the full release of the UK Biobank data (*n* = 487,410), and were processed according to the quality control pipeline (Bycroft et al. 2018). Standard genotype quality control criteria were used (Coleman et al. 2019). Thresholds were variants with a minor allele frequency > 1%, directly genotyped or imputed (IMPUTE INFO metric > 0.4 (McCarthy et al. 2016)). For individuals, a genotype call rate > 98%, concordant phenotypic and genetic gender information, removing third-degree relatives and closer (Manichaikul et al. 2010). Our analyses were limited to individuals of European ancestry due to insufficient numbers of other ancestry groups.

Pregnant females were removed as well as potential outliers in the dataset using lower and upper cutoff limits to each estimated nutrient. These cutoffs were identified by generating scatterplots. We included all time points each participant answered the questionnaire, but restricted our analyses to individuals with complete data on variables that may potentially influence nutrient intake, such as socioeconomic status (SES) and educational attainment (EA). Our sample after exclusion consisted of 350,339 data entries for each estimated nutrient intake covering 163,619 participants (i.e., 77.5% of the original sample). Data cleaning was performed in R version 3.5.3.

### Nutritional intake data

Nutrient intakes (Category 100098) were pre-calculated by the UK Biobank (see Supplementary Materials). We excluded one of each pair of nutrients with correlations >0.7 (Supplementary Figure 1). Accordingly, energy, total sugar, starch, saturated and polyunsaturated fat, magnesium, potassium and retinol were not analysed. These estimates were excluded, except magnesium, due to their high correlation with the key macronutrients, carbohydrates, proteins and fats. There was a high correlation between iron and magnesium intake. We kept iron because iron deficiency is the most prevalent nutritional deficiency and a potential risk factor of psychiatric disorders (Chen et al. 2013). In total, we studied the intake of 14 nutrients - protein, carbohydrates, fats, fibre, food weight, folate, calcium, carotene, iron and vitamins B12, B6, C, D and E - and alcohol.

**Figure 1:**
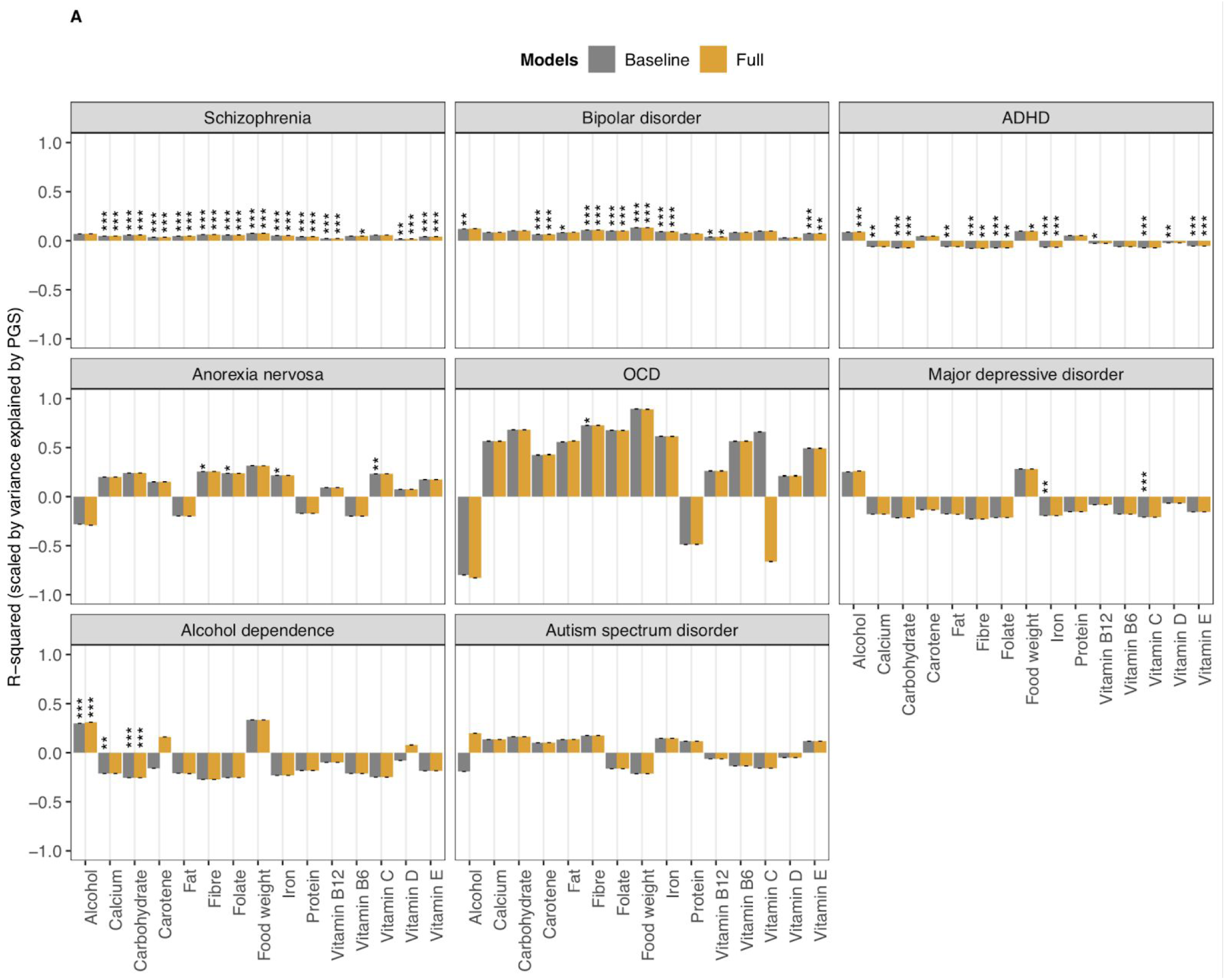
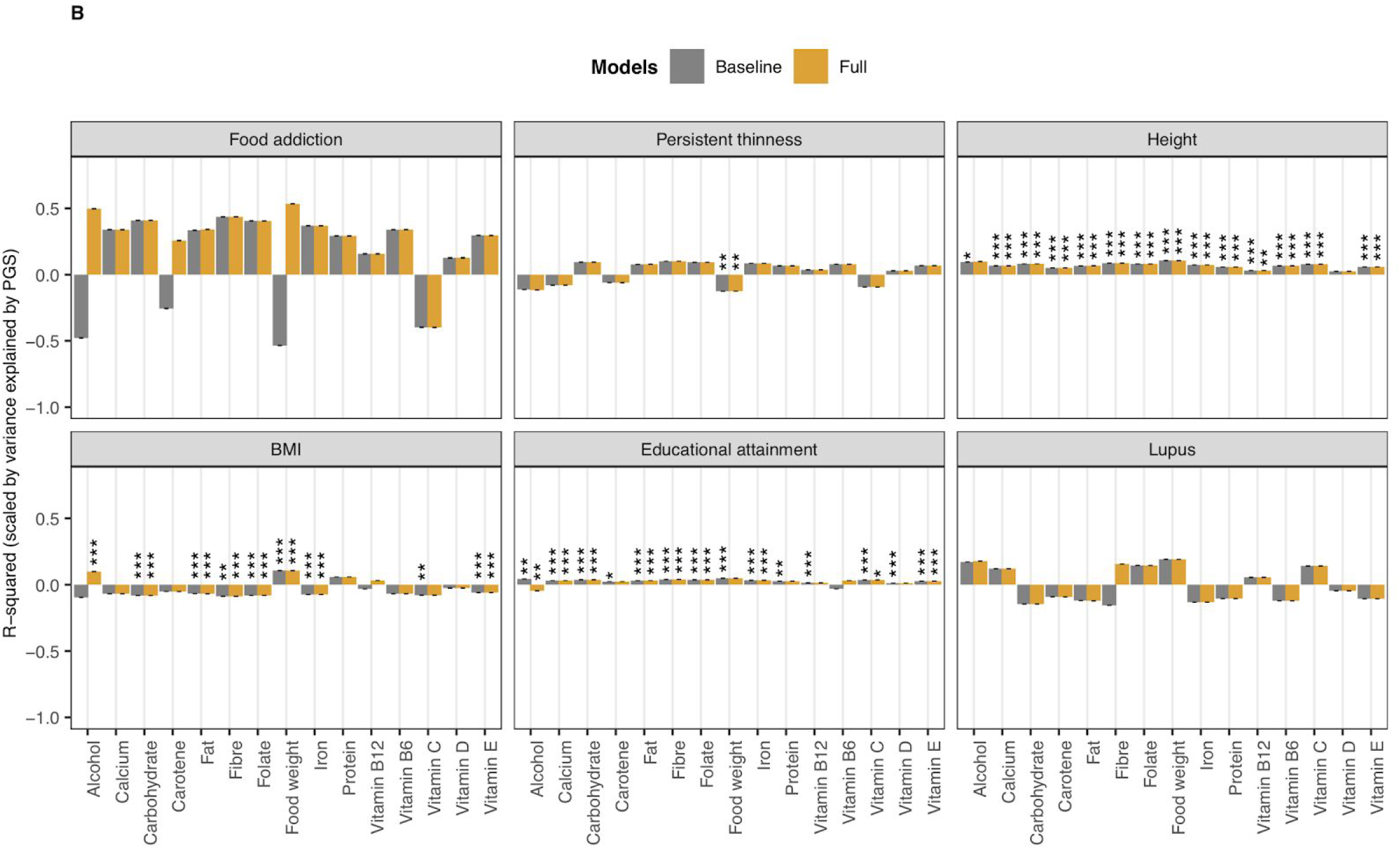
Associations between polygenic scores for psychiatric, behavioural and anthropometric traits and nutrient intake. A) psychiatric disorders and B) psychiatric-related traits, height, BMI, educational attainment and lupus (i.e., negative control). Results are shown from linear mixed-effects model analyses. Y axis shows the R^2^ estimates which have been scaled by the variance explained by the polygenic score predicting itself on the liability scale and have been multiplied by the direction of the coefficient estimate. Colours represent the different models: grey for model 0 (i.e. baseline model) and yellow for model 5 (i.e. full model) in configuration 1. Error bars represent standard errors and asterisks indicate statistically significant estimates. Bonferroni-corrected p value thresholds: ∗ = p < 0.05/132, ∗∗ = p < 0.01/132, ∗∗∗ = p < 0.001/132 Data are represented for 163,619 participants of the UK Biobank cohort The full results can be found in Supplementary Figures 3 and 4

### Polygenic scores

Polygenic scores for psychiatric disorders were constructed for each UK Biobank participant using PRSice version 2.2.1 (Choi and O’Reilly 2019). Single-nucleotide polymorphism (SNP) weights were based on the output from genome-wide association studies (GWAS) of each trait excluding UK Biobank participants (Table 1). We also investigated polygenic scores for BMI, body fat percentage, height, and persistent thinness as body composition is associated with nutrient intake. The latter to compare low BMI with anorexia nervosa - both have low BMI in common, but they differ in their psychiatric symptoms (Hübel et al. 2020). We included polygenic scores for educational attainment, because it has a negative genetic correlation with body composition (Hübel et al. 2019), and lupus as a negative control, because we expected it not to be associated with nutrient intake. Finally, a polygenic score for food addiction was included to investigate what components of food intake may promote an addictive-like response in individuals. To minimise multiple testing, we selected SNPs with P-values <0.2 into the scores.

**Table 1:** Table summarising genome-wide association study (GWAS) discovery sample size, SNP-based heritability on the observed scale

| Trait | GWAS sample | Observed scale SNP-based heritability |
| --- | --- | --- |
| Anorexia nervosa (Watson et al. 2019) | 16,224 cases and 52,460 controls | 17.4% (1.2%) |
| Obsessive-compulsive disorder (International Obsessive Compulsive Disorder Foundation Genetics Collaborative (IOCDF-GC) and OCD Collaborative Genetics Association Studies (OC GAS) 2018) | 2,688 cases and 7,037 controls | 33.8% (4.8%) |
| Educational attainment (Lee et al. 2018) | 766,345 individuals | 10.7% (0.2%) |
| Schizophrenia (Schizophrenia Working Group of the Psychiatric Genomics Consortium 2014) | 33,610 cases and 43,456 controls | 45.5% (1.6%) |
| Attention-deficit/hyperactivity disorder (Demontis et al. 2017) | 19,099 cases, 34,194 controls | 24% (1.5%) |
| Alcohol dependence (Walters et al. 2018) | 11,569 cases and 34,999 controls | 5.4% (1%) |
| Major depressive disorder (Wray et al. 2018) | 116,404 cases and 314,990 controls | 5.7% (0.2%) |
| Bipolar disorder (Stahl et al. 2019) | 20,352 cases and 31,358 controls | 21% (1.1%) |
| Autism spectrum disorder (Grove et al. 2019) | 18,381 cases and 27,969 controls | 12% (1.0%) |
| Food addiction (Cornelis et al. 2016) | 9,314 females | 10% (4%) |
| Persistent thinness (Riveros-McKay et al. 2019) | 1,471 cases and 6,460 controls | 16.7% (3.9%) |
| Height (Wood et al. 2014) | 253,288 individuals | 31.2% (1.4%) |
| BMI (Locke et al. 2015) | 322,154 individuals | 13% (0.5%) |
| Body fat percentage (Lu et al. 2016) | 100,716 individuals | 10.4% (0%) |
| Systemic lupus erythematosus (Bentham et al. 2015) | 7,219 cases and 15,991 controls | 33.3% (9.7%) |

### Linear Mixed Effects Models

We adopted a linear mixed-effects modeling approach to determine whether having an underlying genomic risk for a psychiatric disorder influences eating behaviour while accounting for correlations among repeated assessments within an individual (Mallinckrodt, Clark, and David 2001). Specifically, we used the lmerTest package in R (Kuznetsova, Brockhoff, and Christensen 2017) which extends the ‘lmerMod’ class of the lme4 package by providing P-values for tests for fixed effects. We also used the ‘MuMIn’ package (Bartoń 2020) which calculates R-squared values for mixed-effect models.

#### Main analyses

Our baseline mixed-effects model (model 0; Table 2) included the following fixed effects: polygenic score studied, age, sex and the first 6 ancestry principal components (PCs) calculated on the European subsample. The number of time points each participant answered the dietary assessment was also included as a random effect to account for repeated measures. Table 2 summarises the additional fixed effects included in each of the subsequent models. To identify environmental factors having an important influence on associations between polygenic scores for psychiatric disorders and nutrient intake, additional fixed effects were grouped and differently assessed in each model - model 1) typicality and kind of diet followed, model 2) SES and EA, model 3) physical activity and model 4) ill health - with model 5 adjusting for all fixed effects (configuration 1 in Table 2).

**Table 2:**
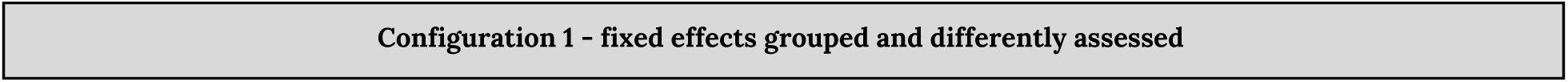
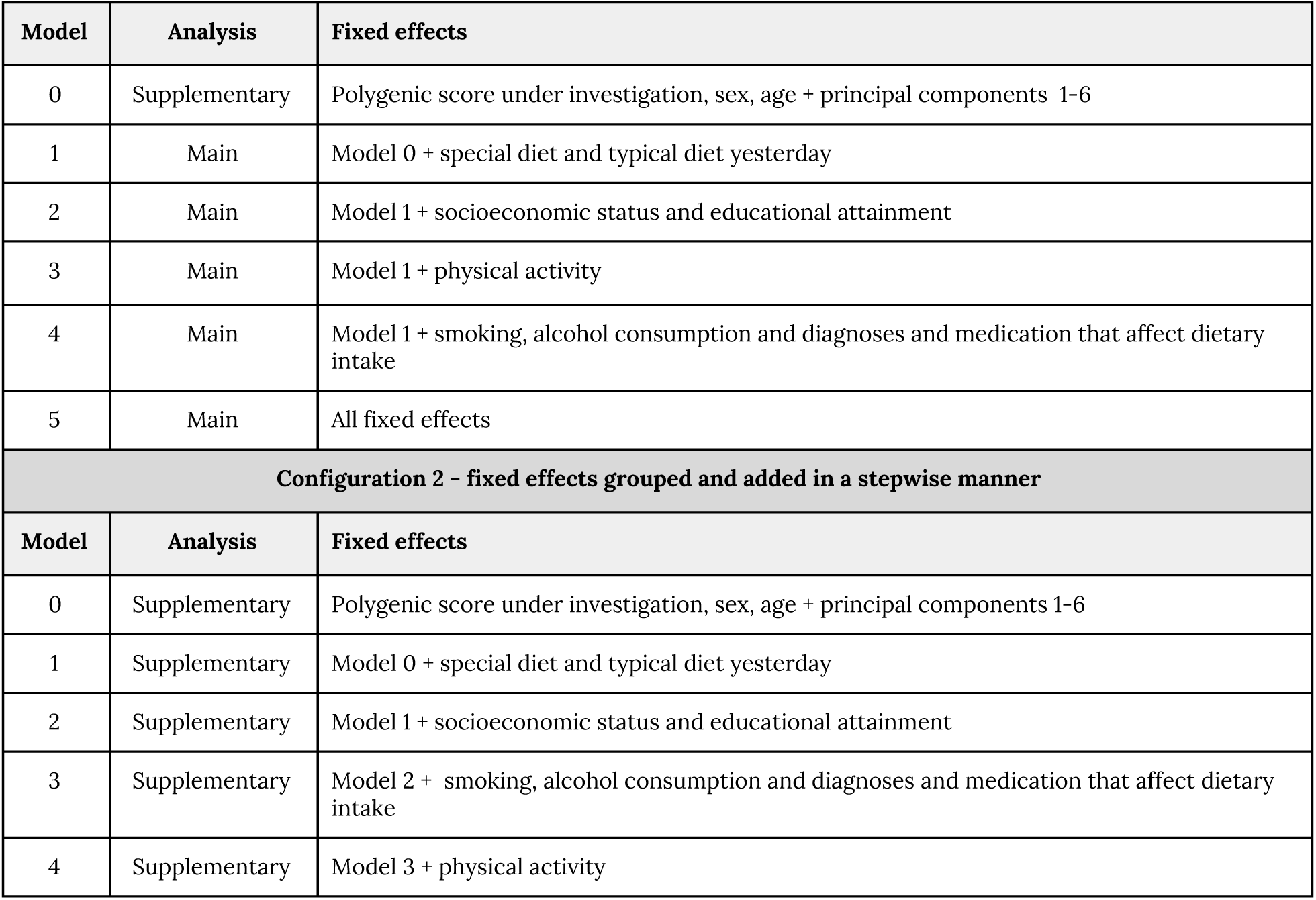
Table summarising the fixed effects included in the linear mixed-effects models. Configurations 1 and 2 represent the models presented in Supplementary Figures 3/5 and 4/6, respectively.

|  |
| --- |
| Configuration 1 - fixed effects grouped and differently assessed |

| Model | Analysis | Fixed effects |
| --- | --- | --- |
| 0 | Supplementary | Polygenic score under investigation, sex, age + principal components 1-6 |
| 1 | Main | Model 0 + special diet and typical diet yesterday |
| 2 | Main | Model 1 + socioeconomic status and educational attainment |
| 3 | Main | Model 1 + physical activity |
| 4 | Main | Model 1 + smoking, alcohol consumption and diagnoses and medication that affect dietary intake |
| 5 | Main | All fixed effects |
| **Configuration 2 - fixed effects grouped and added in a stepwise manner** |  |  |
| Model | Analysis | Fixed effects |
| 0 | Supplementary | Polygenic score under investigation, sex, age + principal components 1-6 |
| 1 | Supplementary | Model 0 + special diet and typical diet yesterday |
| 2 | Supplementary | Model 1 + socioeconomic status and educational attainment |
| 3 | Supplementary | Model 2 + smoking, alcohol consumption and diagnoses and medication that affect dietary intake |
| 4 | Supplementary | Model 3 + physical activity |

To aid in the interpretation of estimated effect sizes, given varying discovery sample sizes for their derivation, polygenic scores were scaled for graphical presentation by dividing the calculated R^2^ of the variance explained by the ability of each polygenic score to predict itself on the liability scale (see Supplementary Materials for unscaled plots). This allowed us to determine the relative importance of each psychiatric disorder and behaviour trait for each nutrient intake, given that some disorders and traits (e.g. schizophrenia, height) have more powerful polygenic scores available. We obtained bootstrapped standard errors (SE) for the R^2^ statistics using the ‘boot’ package in R, with 100 replications.

#### Supplementary/sensitivity analyses

We tested for potential collider bias by including Model 0 (Table 2) to see if an association between the exposure (polygenic score) and outcome (nutrient) exists before adding additional fixed effects. In addition, we present an alternative set of models whereby fixed effects were added in a stepwise manner as opposed to being differentially assessed (Configuration 2 in Table 2), to see whether patterns of association between polygenic scores for psychiatric disorders and nutrient intake are affected by different approaches to add fixed effects.

### Multiple comparisons

Multiple testing correction was performed using matrix decomposition of the correlation matrix of all traits studied (anorexia nervosa, ADHD, OCD, schizophrenia, MDD, alcohol dependence, persistent thinness, food addiction, height, BMI, educational attainment, lupus, alcohol, protein, carbohydrate, fat, fibre, food weight, folate, calcium, carotene, iron and vitamins B12, B6, C, D and E) to identify the number of independent tests to adjust the p-value threshold using Bonferroni correction (not considering supplementary analyses; see Supplementary Materials).

### Data availability

Authors had full access to the data supporting the findings of this study. UK Biobank is an open access resource. Data are available to bona fide scientists, undertaking health-related research that is in the public good. All individual-level data from UK Biobank can be accessed by applying to the UK Biobank Central Access Committee (http://www.ukbiobank.ac.uk/register-apply/).

### Code availability

Analysis code for PRSice can be accessed at https://choishingwan.github.io/PRSice/. Unreported custom code can be made available upon request.

## Results

### Descriptives

Characteristics of the study participants are summarised in Table 3. From our sample of 163,619 participants, 73,853 (45.14%) were males and 89,799 (54.84%) were females. Participants were aged between 40 and 72 (mean age of 56). Forty percent (*n* = 65,296) of our sample completed the questionnaire once, whilst only 2% (*n* = 3,798) completed it on all five occasions. Table 3 also reports the mean, standard deviation (SD), and range for each nutrient. For example, the mean intake of iron was 13.50 mg (SD = 4.67 mg), with a maximum intake of 39.93 mg per day. Supplementary Figure 2 provides a graphical representation of the distribution of each nutrient.

**Table 3:** Descriptive statistics: age, sex, number of questionnaires completed and nutrient intakes in 163,619 participants of the UK Biobank cohort.

|  |  |  |  |
| --- | --- | --- | --- |
| Age |  | 56.22 (7.89) | 40 - 72 |
|  |  | **n (%)** |  |
| Gender | Males | 73,853 (45.1%) |  |
|  | Females | 89,766 (54.9%) |  |
|  | **Total** | **163,619** |  |
| Number of questionnaires completed | 1 | 65,296 (39.9%) |  |
|  | 2 | 39,254 (24%) |  |
|  | 3 | 33,539 (20.5 %) |  |
|  | 4 | 21,732 (13.3%) |  |
|  | 5 | 3,798 (2.3%) |  |

| Nutrient intakes (units) | Mean (SD) | Range |
| --- | --- | --- |
| Alcohol (g) | 16.18 (22.73) | 0 – 149.63 |
| Protein (g) | 80.93 (29.96) | 0 – 294.54 |
| Carbohydrates (g) | 249.78 (87.66) | 0 – 834.88 |
| Fat (g) | 76.19 (31.97) | 0 – 270.35 |
| Fibre (g) | 16.32 (7.02) | 0 – 59.3 |
| Food weight (g) | 3162.84 (808.55) | 0 – 6361.5 |
| Folate (µg) | 298.17 (118.65) | 0 – 995.36 |
| Calcium (mg) | 961.25 (386.64) | 0 – 3934.24 |
| Carotene (µg) | 3044.85 (2860.47) | 0 – 24992.8 |
| Iron (mg) | 13.50 (4.67) | 0 – 39.93 |
| Vitamin B6 (µg) | 2.15 (0.78) | 0 – 6 |
| Vitamin B12 (µg) | 6.13 (4.73) | 0 – 49.33 |
| Vitamin C (µg) | 150.14 (110.63) | 0 – 997.45 |
| Vitamin D (µg) | 2.81 (3.22) | 0 – 24.76 |
| Vitamin E (µg) | 9.13 (4.77) | 0 – 47.31 |

Note: Values are mean (standard deviation) and range (min – max) for age and nutrient intakes, and n (%) for sex and number of assessments completed

### Association of polygenic scores for psychiatric traits with nutrient intake

In the mixed-effects regression analyses of the nutrient intake on polygenic scores of psychiatric disorders and behavioural traits, differential associations emerged (Figure 1). Figure 1 shows the estimated R^2^ values, scaled by the variance explained by the polygenic score predicting itself on the liability scale, for model 0 (i.e. baseline model) and model 5 (i.e. full model) in configuration 1 (See supplementary Figures 3 and 4 for scaled plots, and Supplementary Figures 5 and 6 for unscaled plots, for model configurations 1 and 2, respectively). We found no association between nutrient intake and polygenic scores for lupus (i.e., negative control, as expected), autism spectrum disorder and food addiction.

#### Schizophrenia and bipolar disorder

Polygenic scores for schizophrenia and bipolar disorder showed the strongest associations with nutrient intake (Supplementary Figure 5). The schizophrenia score was modestly but positively associated with most nutrients with the exception of alcohol and vitamin C. We found a positive association between schizophrenia polygenic score and vitamin B6 but only in the full model (model 5 in Supplementary Figure 3/5 or model 4 in Supplementary Figure 4/6). A one SD higher bipolar disorder polygenic score was associated with higher average daily food weight (11.4 g) and a higher average daily intake of alcohol (0.20 g), fat (0.24 g), fibre (0.11 g), folate (1.45 μg), carotene (27.6 μg), iron (0.06 mg), vitamin B12 (0.03 μg) and vitamin E (0.05 μg) (Model 0 in Supplementary Table 1/2), calcium (effect size after adjusting for ill health = 2.94 mg), vitamin C (0.94 μg) and vitamin D (0.02 μg) after adjusting for either the typicality and kind of diet followed, physical activity or ill health which included negative health behaviours and appetite-modulating medication (Supplementary Figure 3/5 and Supplementary Table 1). After adjusting for phenotypic SES and EA the associations between the bipolar disorder polygenic score and alcohol, fat and the vitamins C, D and B12 attenuated and did not remain significant (Supplementary Figure 3/5). We also observed no association between the bipolar disorder polygenic score and fat when fixed effects were included in a stepwise method (Supplementary Figure 4/6).

#### Anorexia nervosa and OCD

A one SD higher anorexia nervosa polygenic score was associated with higher intake of fibre (0.06 g), folate (0.93 μg), iron (0.03 mg) and vitamin C (0.92 μg; Figure 1; Model 0 in Supplementary Table 1/2). For fibre, folate, and iron, these associations were not significant after adjusting for SES, EA, and physical activity, as well as ill health for iron - irrespective of modelling approach (Supplementary Figures 3/5 and 4/6). Similarly, a higher OCD polygenic score was associated with 0.06g higher fibre intake (Model 0 in Supplementary Table 1/2), attenuating when adjusted for EA and SES. Interestingly, OCD showed the greatest magnitude of effect on nutrient intake, proportional to the power of the polygenic score (Figure 1; Supplementary Figure 3 and 4).

#### MDD

A higher polygenic score for MDD was associated with lower intake of iron (−0.04 mg) and vitamin C (−1.13 μg; Figure 1; Model 0 in Supplementary Table 1/2). We also observed a positive association with alcohol intake and negative association with vitamin E after adjusting for physical activity. We found the association between MDD polygenic score and iron attenuated when adjusting for SES and EA or ill health (Supplementary Figures 3/5 and 3/5). In addition, stepwise inclusion of additional fixed effects attenuated the association between MDD polygenic score and vitamin C (Supplementary Figure 3/5).

#### ADHD and alcohol dependence

A one SD higher ADHD polygenic score was associated with 0.23 g higher alcohol consumption and a 7.6 g higher overall food weight (Figure 1). These associations were not significant after adjusting for physical activity, as well as ill health (Supplementary Figure 3/5). However, they remained significant when fixed effects were adjusted for in a stepwise manner (Model 4 in Supplementary Figure 4/6). A one SD higher ADHD polygenic score was also associated with lower intake of carbohydrates (−1.36 g), fat (−0.29 g), fibre (−0.08 g), folate (−1.48 μg), calcium (−3.45 mg), iron (−0.09 mg), and vitamins B12 (−0.03 μg), C (−1.26 μg), D (−0.02 μg) and E (−0.09 μg) (Model 0 in Supplementary Table 1/2). However, associations with fat, calcium, vitamin C and vitamin D were not significant after adjusting for SES and EA, as well as physical activity for fat and vitamin B12, and ill health for vitamin D - irrespective of whether fixed effects were grouped or adjusted for in a stepwise approach. Similarly to ADHD, a higher alcohol dependence polygenic score was associated with 0.4 g higher alcohol intake and 5.9 g higher food weight (Figure 1; Model 0 in Supplementary Table 1/2), the latter only when adjusting for SES and EA. The association between alcohol dependence polygenic score and alcohol intake was dependent of SES and EA, and reduced when adjusting for alcohol consumption (as expected, Supplementary Figures 3/5 and 4/6). We also found negative associations with the intake of carbohydrates (−1.18 g) and calcium (−3.42 mg) (Model 0 in Supplementary Table 1/2). The latter was not significant after adjusting for ill health.

#### Height, BMI and persistent thinness

We also studied polygenic scores for height, BMI and persistent thinness. As with schizophrenia, the height polygenic score was positively associated with all nutrients, except vitamin D. The association between the height polygenic score and alcohol was attenuated by adjusting for SES and EA (Supplementary Figure 3/5 and 4/6). In addition, a one SD higher BMI polygenic score was associated with a 0.2 mg higher alcohol, and a 9.67 g higher food weight, but with lower intake of carbohydrates (−1.90 g), fats (−0.58 g) and fibre (−0.07 g) (Model 0 in Supplementary Table 1/2). The association between the BMI polygenic score and alcohol intake was restricted to those models adjusting for ill health (i.e. models 4 and 5 in Supplementary Figure 3/5 or models 3 and 4 in Supplementary Figure 4/6). As a sensitivity analysis, we report similar findings for a body fat percentage polygenic score (Supplementary Figure 7). Furthermore, we found a one SD higher polygenic score for persistent thinness was associated with 8.61g lower food weight (Model 0 in Supplementary Table 1/2).

#### Educational attainment

Finally, we found a one SD higher polygenic score for educational attainment was positively associated with all nutrients, except vitamin B6. Adjusting for ill health reversed the association between the education attainment polygenic score and alcohol intake (0.20 g in model 0 compared to −0.28 g in model 4 ill health adjustment - Supplementary Table 1). In addition, we found adjustment for phenotypic SES/EA weakened associations, with protein, food weight, carotene and vitamin B12 not meeting our significance threshold anymore (Supplementary Figure 3/5).

## Discussion

A recent UK Biobank study found significant genetic correlations between schizophrenia and two diet groups, one representing a meat-related diet and the other a fish and plant-related diet (Niarchou et al. 2020). This spurred our in-depth investigation of the association between polygenic scores for psychiatric disorders and nutrient intake. We show for the first time that polygenic scores for seven psychiatric disorders and several behavioral and anthropometric traits show significant associations with self-reported nutrient intake on an average day.

Polygenic scores for schizophrenia, bipolar disorder and ADHD showed the highest number of significant associations with the intake of specific nutrients. However, scaling the calculated R^2^ values by their power (the variance explained by the polygenic score predicting itself on the liability scale) revealed that the OCD polygenic score had the greatest magnitude of effect on nutrient intake. Given that the OCD GWAS was relatively small compared to the other psychiatric disorder GWAS, these findings suggest that with larger and more powerful GWAS of OCD associations of OCD with diet traits may become more apparent.

We also investigated polygenic scores for anorexia nervosa and persistent thinness. Both are characterised by low BMI but differ in their psychiatric symptoms (Hübel et al. 2020). Individuals with persistent thinness do not suffer from undernutrition or exhibit any typical clinical features such as amenorrhea, fear of weight gain, or hormonal abnormalities commonly seen in anorexia nervosa (Estour et al. 2017). Notably, polygenic scores for anorexia nervosa and persistent thinness had distinct associations with nutrient intake. The anorexia nervosa polygenic score was associated with higher intake of fibre, folate, iron and vitamin C. In contrast, a higher persistent thinness polygenic score was associated with lower food weight but nothing else. These findings complement the clinical definition of persistent thinness and support the suggestion that anorexia nervosa and persistent thinness may be genetically distinct (Riveros-McKay et al. 2019).

Polygenic scores for ADHD and alcohol dependence had a similar pattern of association with nutrient intake, both being associated with higher alcohol intake. Given that impulsivity is common among individuals with ADHD and those who are alcohol dependent (Miller et al. 2010; Dick et al. 2010), alcohol use may relate to an individual’s impulsive tendency to use alcohol for the immediate reward associated with drinking (Shin, Hong, and Jeon 2012). The association between genetic risk for alcohol dependence and alcohol intake was not significantly influenced by phenotypic SES and EA. This is interesting because SES and EA have been identified as prominent risk factors for alcohol dependence (Collins 2016; Springer 2019). However, our findings suggest that other unaccounted risk factors, such as peer pressure (Dumas, Ellis, and Wolfe 2012) and living in a family or culture where frequent alcohol use/abuse is accepted (Lo Monaco et al. 2020), may be more influential in an individual’s genetic liability to develop an alcohol problem.

A higher educational attainment polygenic score was positively associated with nutrient intake, providing a genetic basis for the phenotypic association between EA and dietary intake (Rippin et al. 2020). Adjusted for phenotypic EA attenuated associations, as expected. However, several remained significant including associations with carbohydrate and fat intake. This could reflect genetic pleiotropy because a recent study found that the EA polygenic score captures DNA variants shared between educational achievement and personality traits, including agreeableness, openness, conscientiousness, and academic motivation (Smith-Woolley, Selzam, and Plomin 2019). Furthermore, personality traits have previously been linked to taste preference and eating behaviour (Keller and Siegrist 2015). This offers a potential explanation for the significant associations between EA polygenic score and specific nutrients after adjusting for phenotypic EA.

Contrary to our expectations, a higher BMI polygenic score was associated with lower intake of carbohydrates, fats and fibre. We observed similar findings for a body fat percentage polygenic score. This suggests that the higher BMI in obese individuals may not originate from a biological liability for higher fat and carbohydrate intake but may be associated with other factors such as dysfunction of autonomic neural circuits (Turnbaugh et al. 2006). Alternatively, underreporting bias may have occurred because individuals with a higher BMI polygenic score reported higher food intake, when measured as weight, but lower intake of specific nutrients. This misreporting may reflect socially desirable responses and low ability to report own dietary intake (Bel-Serrat et al. 2016). Based on our findings, these socially desirable responses may be a lower reported intake of fat, carbohydrates, and total energy intake than actual intake.

Finally, we grouped fixed effects into distinct groups to determine which environmental factors influence the association between polygenic scores for psychiatric disorders and nutrient intake. Using this approach, we found some evidence for collider bias whereby the exposure and outcome independently cause a third variable, inducing associations where there is no true effect. For example, no association was observed between alcohol dependence polygenic score and food weight until we adjusted for SES and EA. This suggests that environmental factors commonly believed to confound the relationship between psychiatric disorders and dietary behaviour may act as colliders. ‘Future studies should identify potential colliders and their magnitude of effect.

### Limitations

There are several important caveats that need to be taken into account when interpreting these findings. First, dietary intake in the UK Biobank was self-reported, as with most nutritional epidemiology studies, and this method of data collection has inherent limitations (Smith, Jobe, and Mingay 1991; Hebert et al. 1995). We used repeated measures to reduce reporting bias but objective measures of dietary intake, which are currently unavailable at these large sample sizes, would be superior. Furthermore, although our polygenic scores were constructed from the largest available GWAS, some phenotypes still had relatively small sample sizes and consequently these polygenic scores are more weak predictors. We attempted to address this by scaling polygenic scores however these analyses should be repeated when sample sizes have increased.

## Conclusion, implications and future directions

To summarise, polygenic propensity for a psychiatric disorder is associated with nutritional intake. This has important implications for future treatment strategies. Our findings encourage further research into the shared biological pathways and common environmental factors influencing both nutritional intake and psychiatric traits. This could help develop integrative treatments that prevent development of additional comorbidities in individuals with a psychiatric diagnosis.

Future research should explore the developmental association between psychiatric traits and nutritional intake, to capture age-dependent differences. These studies should focus on the impact of SES, EA and physical activity, because these factors influenced several associations observed. In addition, we found having genetic risk for schizophrenia was associated with higher fat intake and food weight even after controlling for antipsychotics. However, mendelian randomisation using GWAS findings suggests schizophrenia is negatively associated with body composition (Hübel et al. 2019). Future work should attempt to identify potential causes underlying this differential association. Apps focused on health and fitness have emerged on the smartphone market: these studies should take advantage of food-tracking apps which may provide a better alternative to dietary recall questionnaires.

## Data Availability

Authors had full access to the data supporting the findings of this study. UK UK Biobank is an open access resource. Data are available to bona fide scientists, undertaking health-related research that is in the public good. All individual-level data from UK Biobank can be accessed by applying to the UK Biobank Central Access Committee.

http://www.ukbiobank.ac.uk/register-apply/

## Acknowledgements

This research has been conducted using the UK Biobank Resource under application number 23395. We are extremely grateful to all the participants who took part in this study, and the whole UK Biobank team..

## Author contributions

All authors substantially contributed to the design of the research. Avina K. Hunjan, Christopher Hübel, and Yuhao Lin provided essential materials, analysed data or performed statistical analyses. Avina K. Hunjan wrote the paper, and all authors drafted the work, approved the final manuscript, and agreed to be accountable for all aspects of the work. Thalia C. Eley and Gerome Breen had primary responsibility for final content.

