## Supplementary Materials for "Association between polygenic propensity for a psychiatric disorder and nutrient intake"

**Recruitment and completion of dietary assessment**

Participants recruited between April 2009 and September 2010 completed the dietary assessment at the assessment centre. After the recruitment period closed, the dietary assessment was completed remotely on four separate occasions (cycle 1, February-April 2011; cycle 2, June-September 2011; cycle 3, October-December 2011; cycle 4, April-June 2012) by participants who provided an email address at recruitment. In total, 211,036 participants completed the dietary assessment at least once.

**Calculation of nutrient intakes**

Nutrient intakes (Category 100098) were pre-calculated by UK Biobank using a three-step process. First, each food and drink type listed in the dietary assessment tool was assigned a portion size, using the ‘Food Portion Sizes’ atlas (12). Secondly, the quantity of each food and drink consumed was calculated by multiplying the assigned portion size by the amount consumed. Using this information, the nutrient intakes for each participant were calculated by multiplying the quantity consumed by the nutrient composition of the food or drink, as taken from a food composition database (13).

**Matrix decomposition to identify number of independent tests**

We used a Bonferroni threshold (by correcting α as α/*N*) to estimate the number of significant genetic correlations, with *N* = estimated number of independent tests. We built a similarity matrix reflecting the trait similarity where *D =* number of principal components accounting for 99.5% of the data variance in the genetic correlation matrix. In that case, *D* is the estimated number of independent traits (GWAS), and the number of independent tests can be computed as *N* = (*D*(*D*-1))/2.

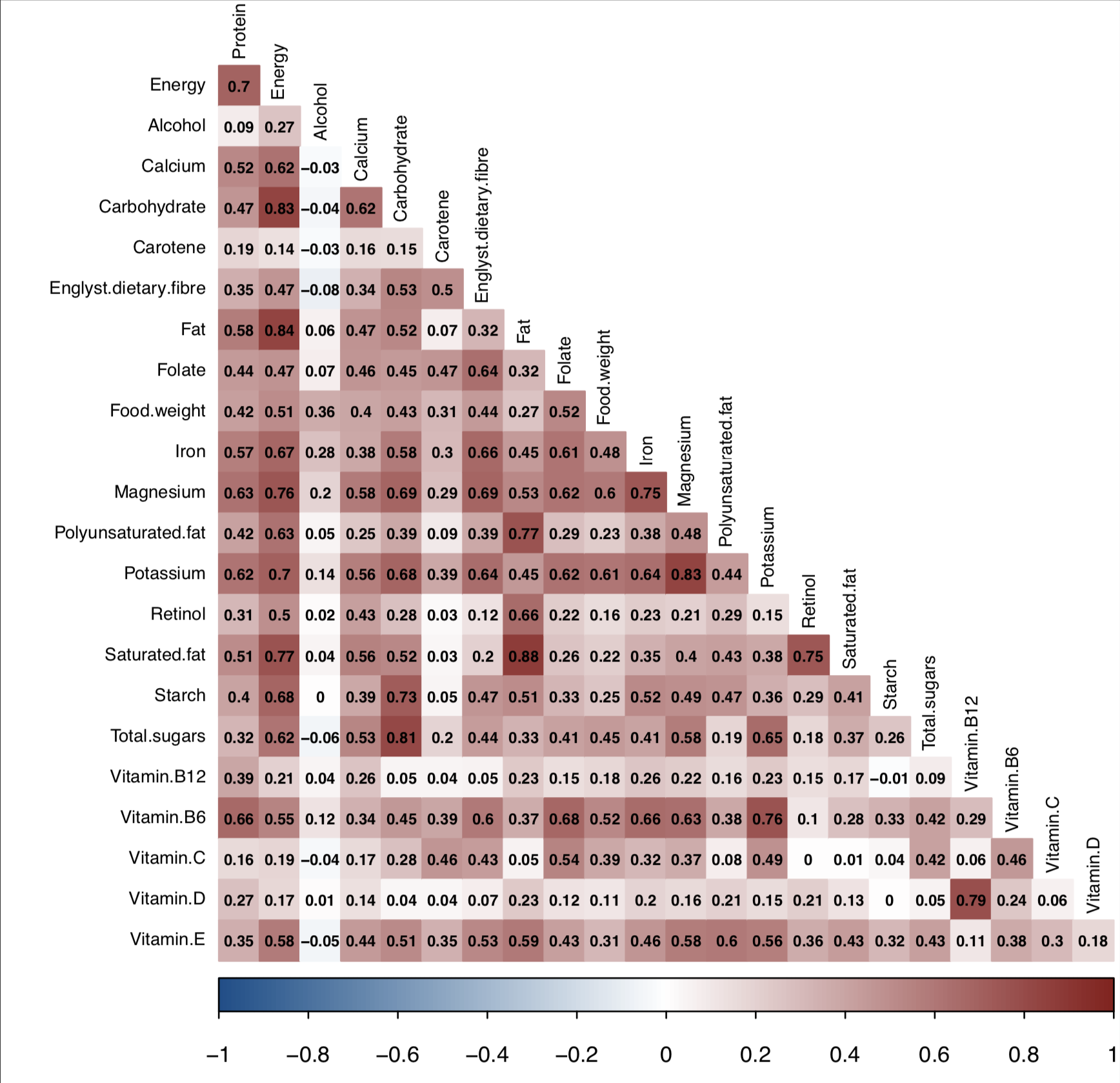

**Supplementary Figure 1:** Correlation matrix of nutrients. For each participant all time points were included for each nutrient. Data are represented for 163,619 participants of the UK Biobank cohort.

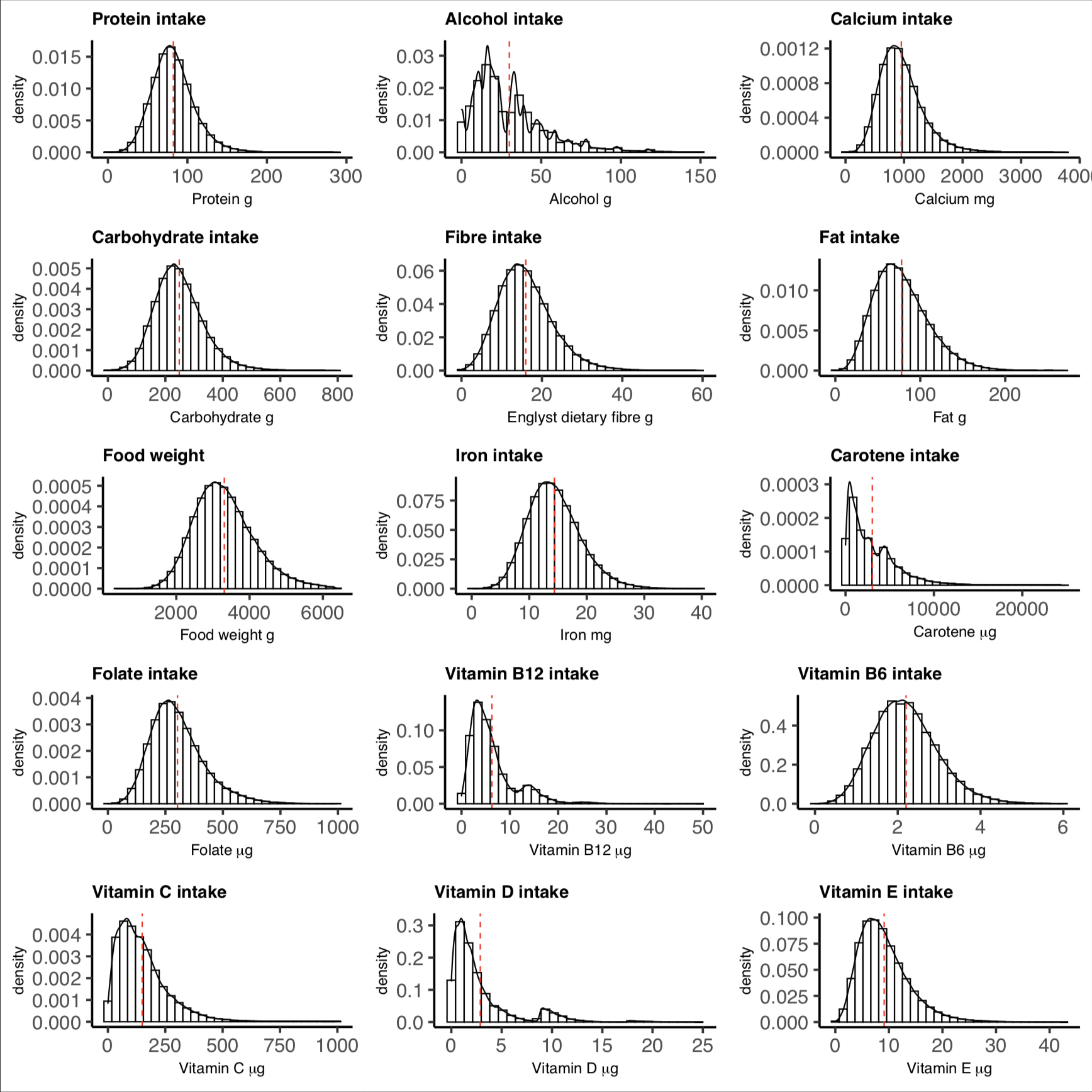

**Supplementary Figure 2:** Histogram plots of each nutrient intake with a fitted normal density curve. For each participant all time points were plotted for each nutrient. The red dashed line represents the mean. Data are represented for 163,619 participants of the UK Biobank cohort.

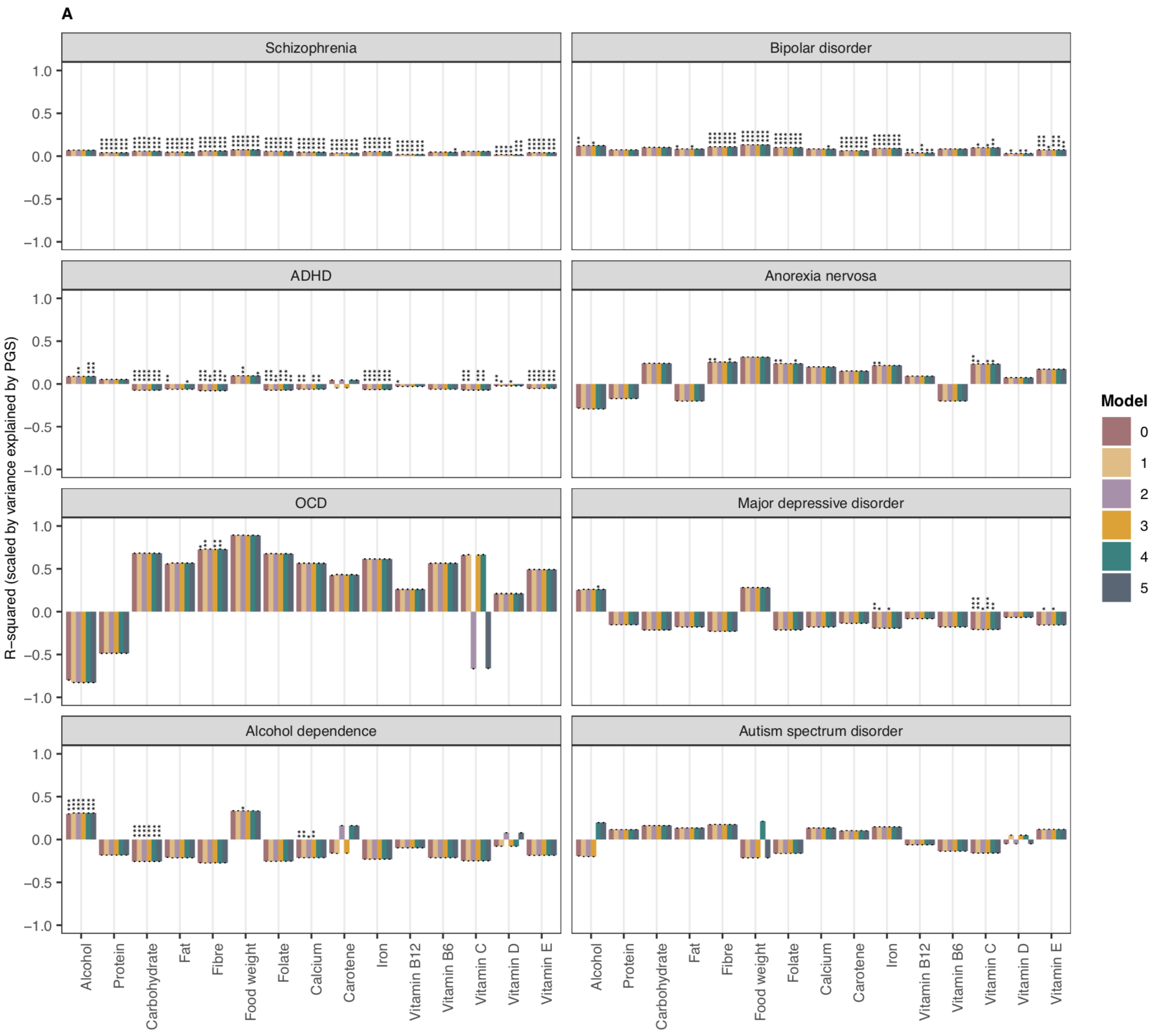

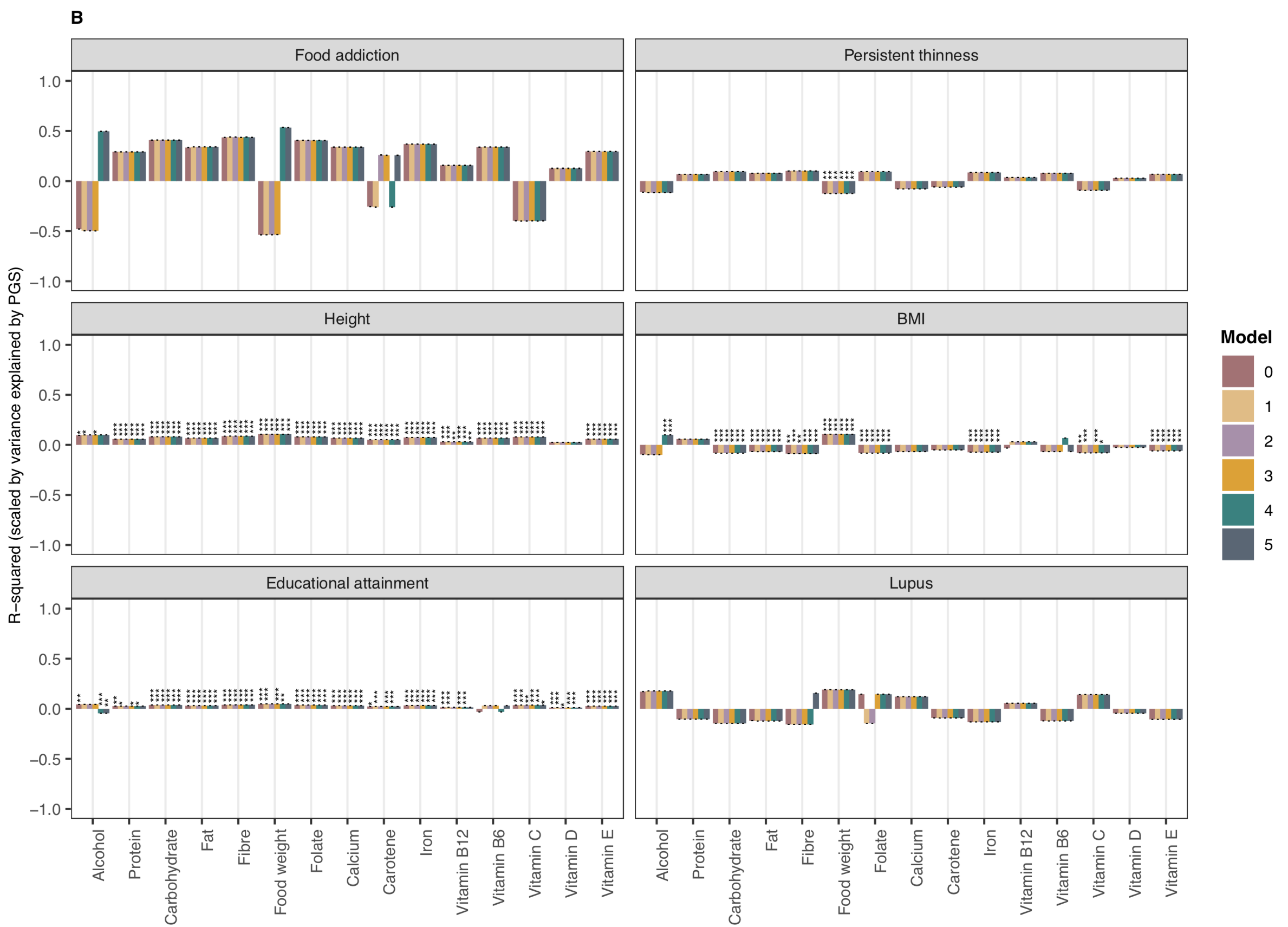

**Supplementary Figure 3:** Scaled associations between polygenic scores for psychiatric, behavioural and anthropometric traits and nutrient intake - configuration 1 from Table 2: A) psychiatric disorders and B) psychiatric-related traits, height, BMI, educational attainment and lupus.

Results are shown from linear mixed-effects model analyses. Y axis shows the R-squared estimates which have been scaled by the variance explained by the PGS predicting itself on the liability scale and have been multiplied by the direction of the coefficient estimate. Colours represent the different models:

Model 0 - Sex, age and PC 1-6

Model 1: Sex, age, PC 1-6, special diet and typical diet yesterday

Model 2: Model 1 + socioeconomic status and educational attainment

Model 3: Model 1 + Physical activity

Model 4: Model 1 + diagnoses and medication that affect food intake, smoking and alcohol consumption

Model 5: All fixed effects

Error bars represent standard errors and asterisks indicate statistically significant estimates. Bonferroni-corrected p value thresholds: ∗ = p < 0.05/132, ∗∗ = p < 0.01/132, ∗∗∗ = p < 0.001/132

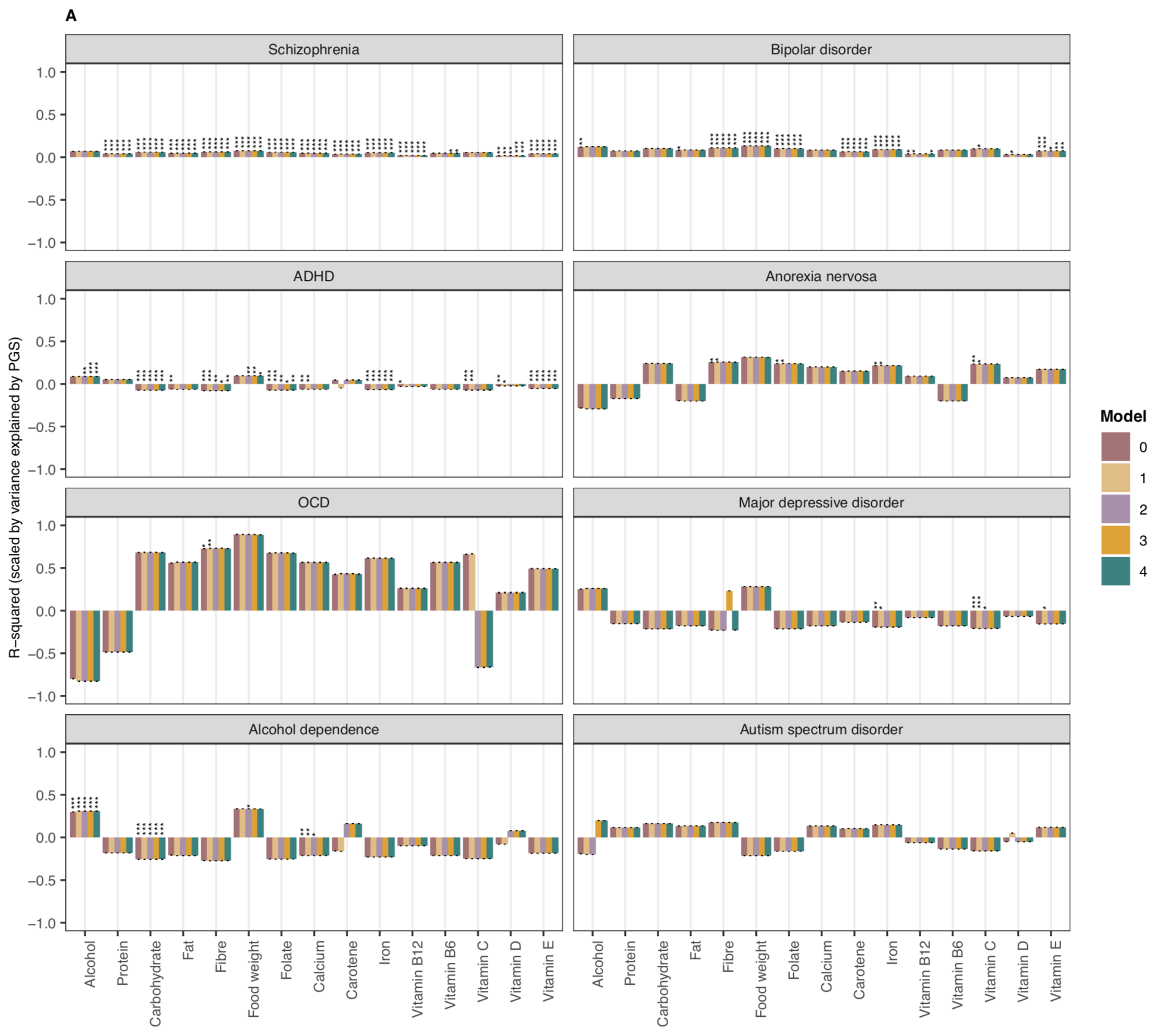

**Supplementary Figure 4:** Scaled associations between polygenic scores for psychiatric, behavioural and anthropometric traits and nutrient intake - configuration 2 from Table 2: A) psychiatric disorders and B) psychiatric-related traits, height, BMI, educational attainment and lupus.
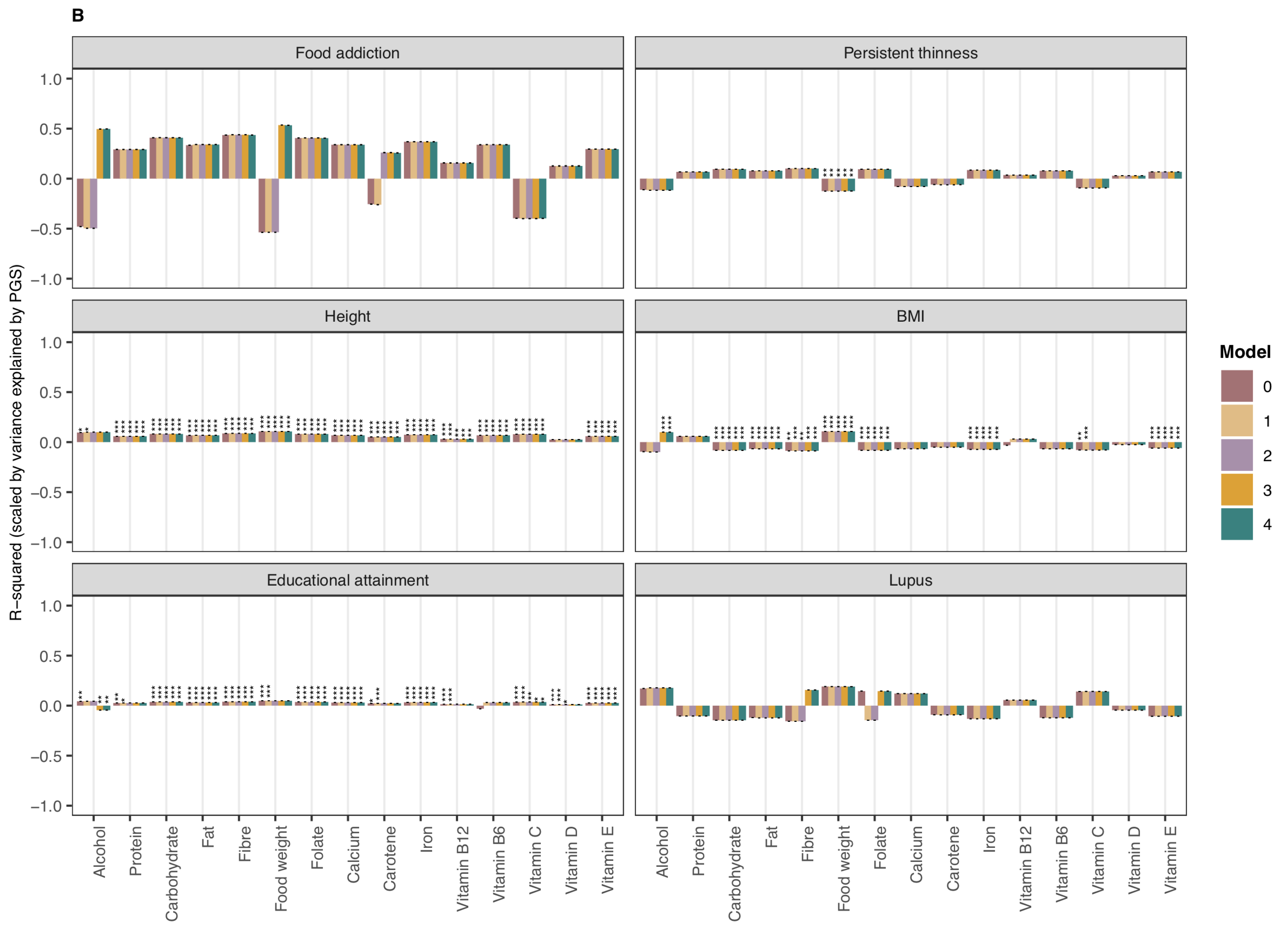

Results are shown from linear mixed-effects model analyses. Y axis shows the R-squared estimates which have been scaled by the variance explained by the PGS predicting itself on the liability scale and have been multiplied by the direction of the coefficient estimate. Colours represent the different models:

Model 0 - Sex, age and PC 1-6

Model 1: Sex, age, PC 1-6, special diet and typical diet yesterday

Model 2: Model 1 + socioeconomic status and educational attainment

Model 3: Model 2 + diagnoses and medication that affect food intake, smoking and alcohol consumption

Model 4: Model 3 + physical activity

Error bars represent standard errors and asterisks indicate statistically significant estimates. Bonferroni-corrected p value thresholds: ∗ = p < 0.05/132, ∗∗ = p < 0.01/132, ∗∗∗ = p < 0.001/132

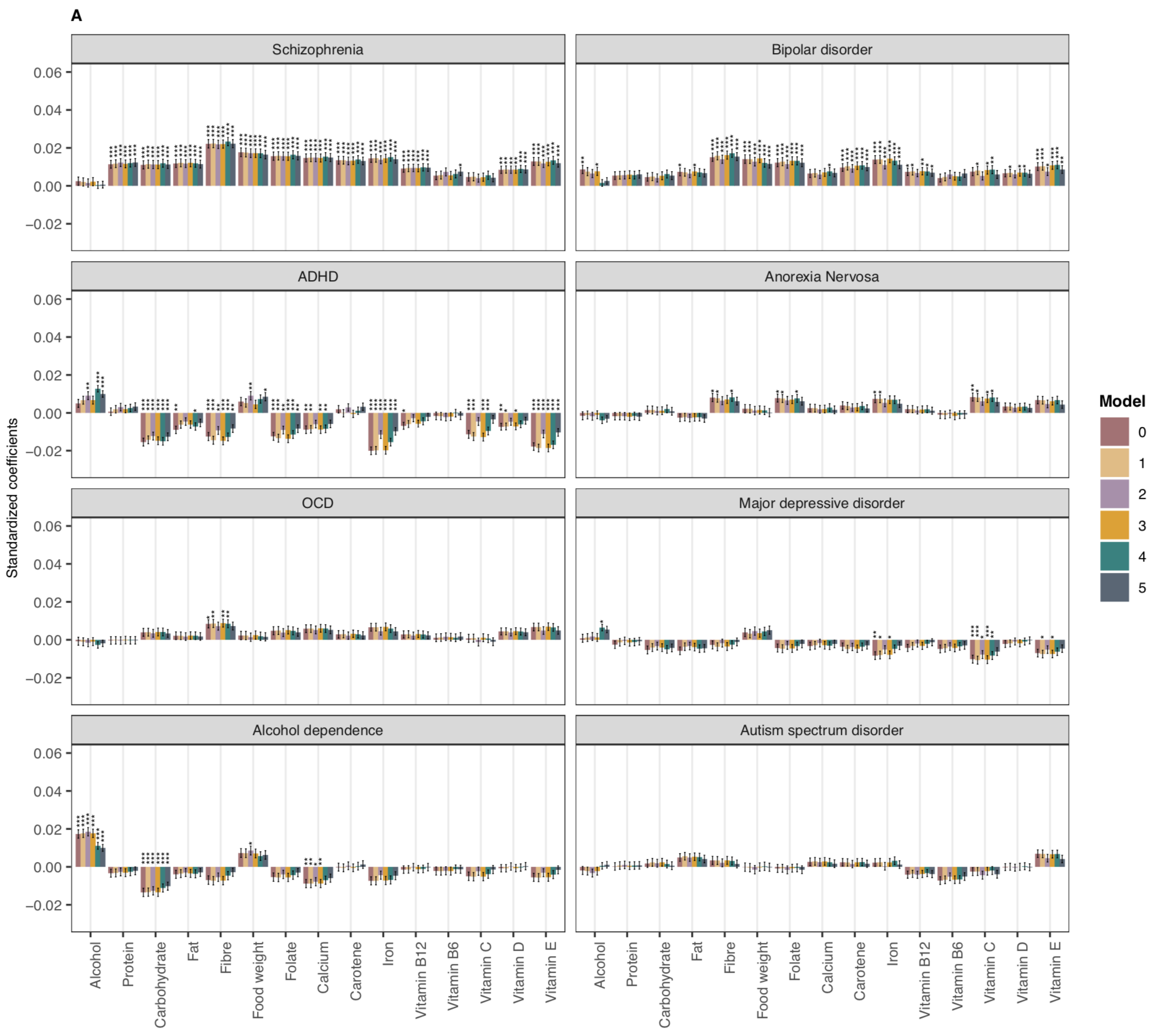

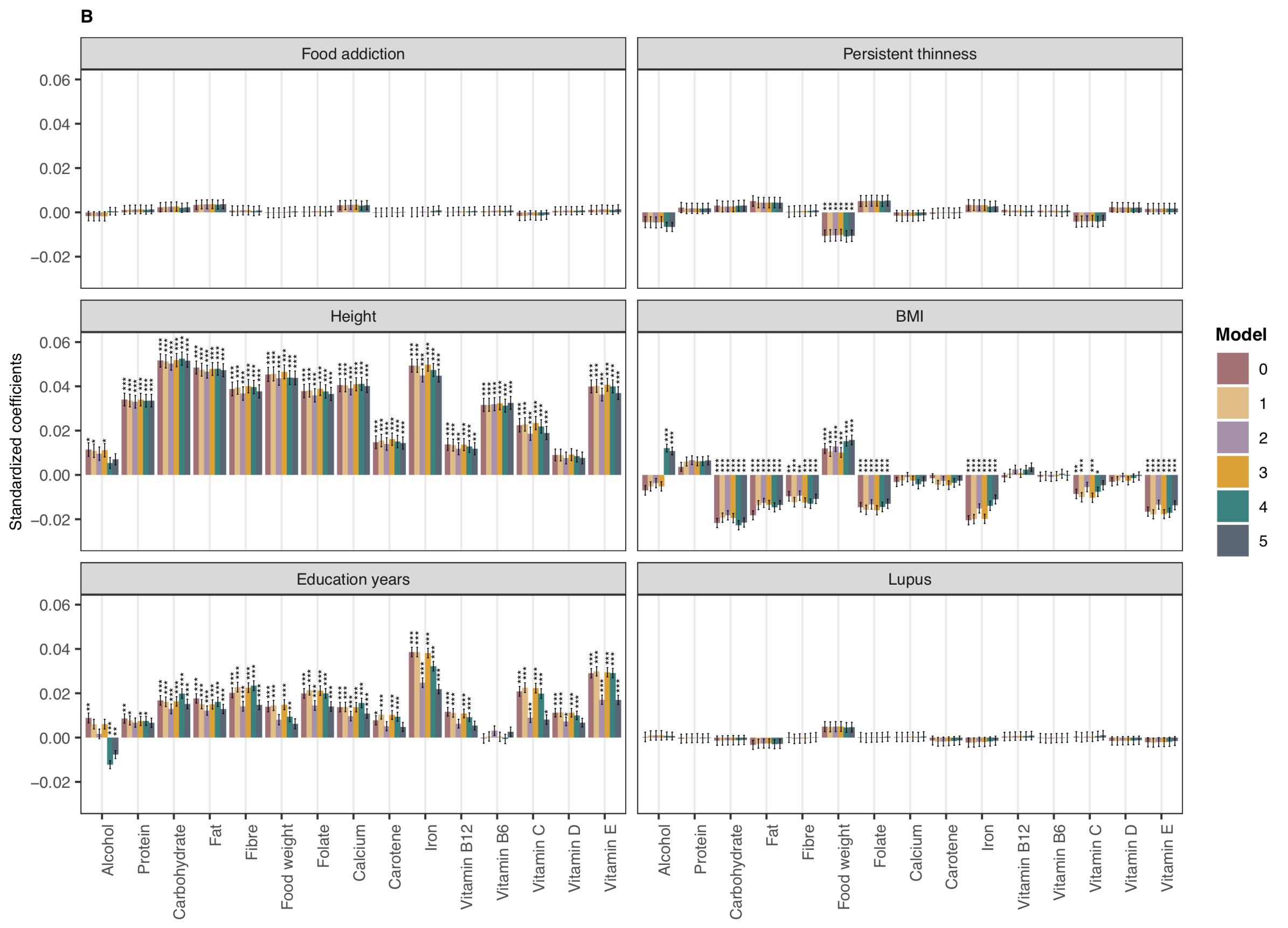

**Supplementary Figure 5:** Unscaled associations between polygenic scores for psychiatric, behavioural and anthropometric traits and nutrient intake - configuration 1 from Table 2. A) psychiatric disorders and B) psychiatric-related traits, height, BMI, education years and lupus .

Results are shown from linear mixed-effects model analyses. Y axis shows the standardised β coefficient estimate (measured in units of standard deviation). PGSs and nutrient intakes were standardised with the figure showing the effect of a one standard deviation increase in PGS on nutrient intake. Colours represent the different models:

Model 0 - Sex, age and PC 1-6

Model 1: Sex, age, PC 1-6, special diet and typical diet yesterday

Model 2: Model 1 + socioeconomic status and educational attainment

Model 3: Model 1 + Physical activity

Model 4: Model 1 + diagnoses and medication that affect food intake, smoking and alcohol consumption

Model 5: All fixed effects

Error bars represent standard errors and asterisks indicate statistically significant estimates. Bonferroni-corrected p value thresholds: ∗ = p < 0.05/132, ∗∗ = p < 0.01/132, ∗∗∗ = p < 0.001/132

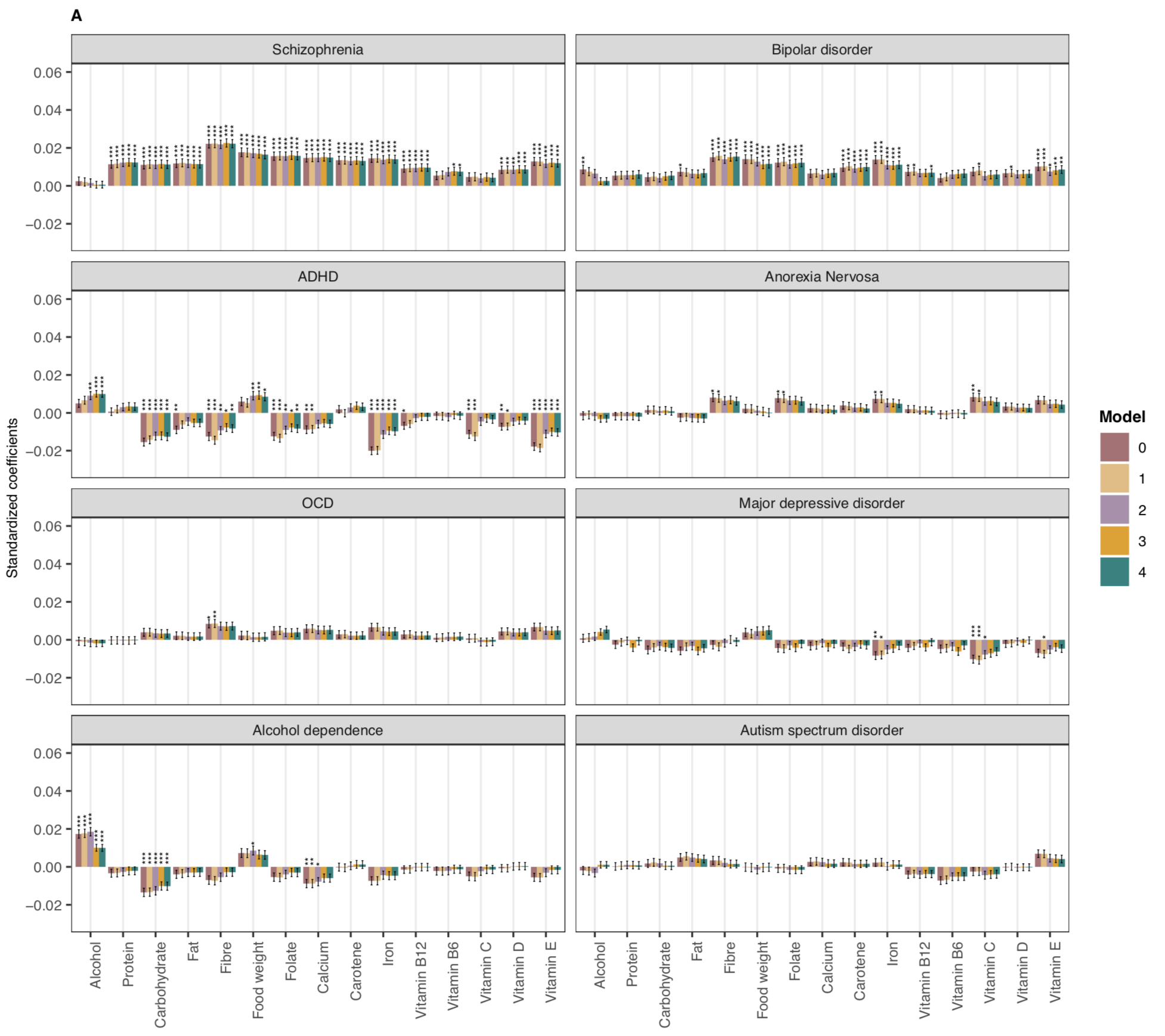

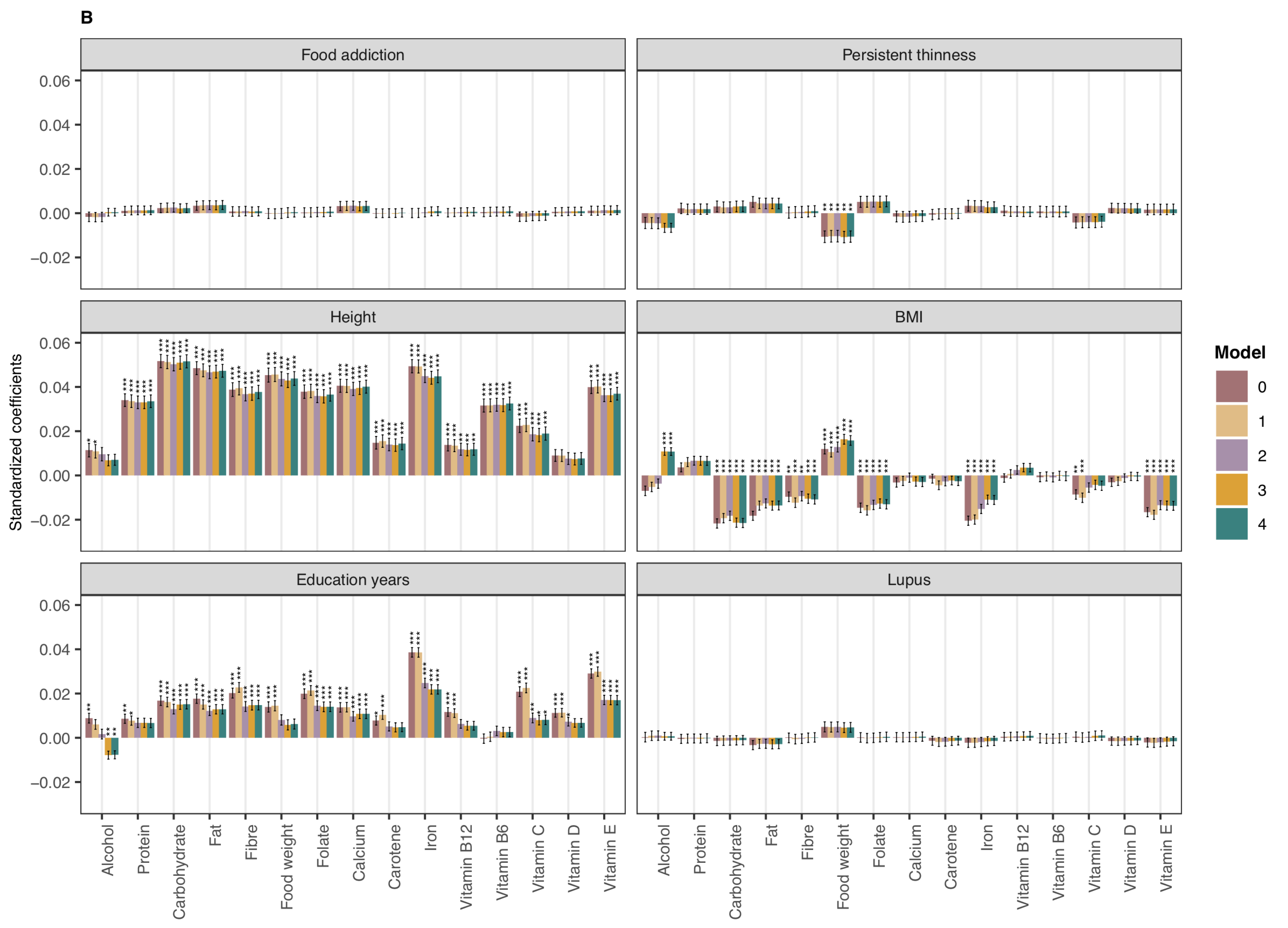

**Supplementary Figure 6:** Unscaled associations between polygenic scores for psychiatric, behavioural and anthropometric traits and nutrient intake - configuration 2 from Table 2. A) psychiatric disorders and B) psychiatric-related traits, height, BMI, education years and lupus .

Results are shown from linear mixed-effects model analyses. Y axis shows the standardised β coefficient estimate (measured in units of standard deviation). PGSs and nutrient intakes were standardised with the figure showing the effect of a one standard deviation increase in PGS on nutrient intake. Colours represent the different models:

Model 0 - Sex, age and PC 1-6

Model 1: Sex, age, PC 1-6, special diet and typical diet yesterday

Model 2: Model 1 + socioeconomic status and educational attainment

Model 3: Model 2 + diagnoses and medication that affect food intake, smoking and alcohol consumption

Model 4: Model 3 + physical activity

Error bars represent standard errors and asterisks indicate statistically significant estimates. Bonferroni-corrected p value thresholds: ∗ = p < 0.05/132, ∗∗ = p < 0.01/132, ∗∗∗ = p < 0.001/132

**Supplementary Figure 7:** Unscaled associations between polygenic scores for BMI and body fat percentage, and nutrient intake.
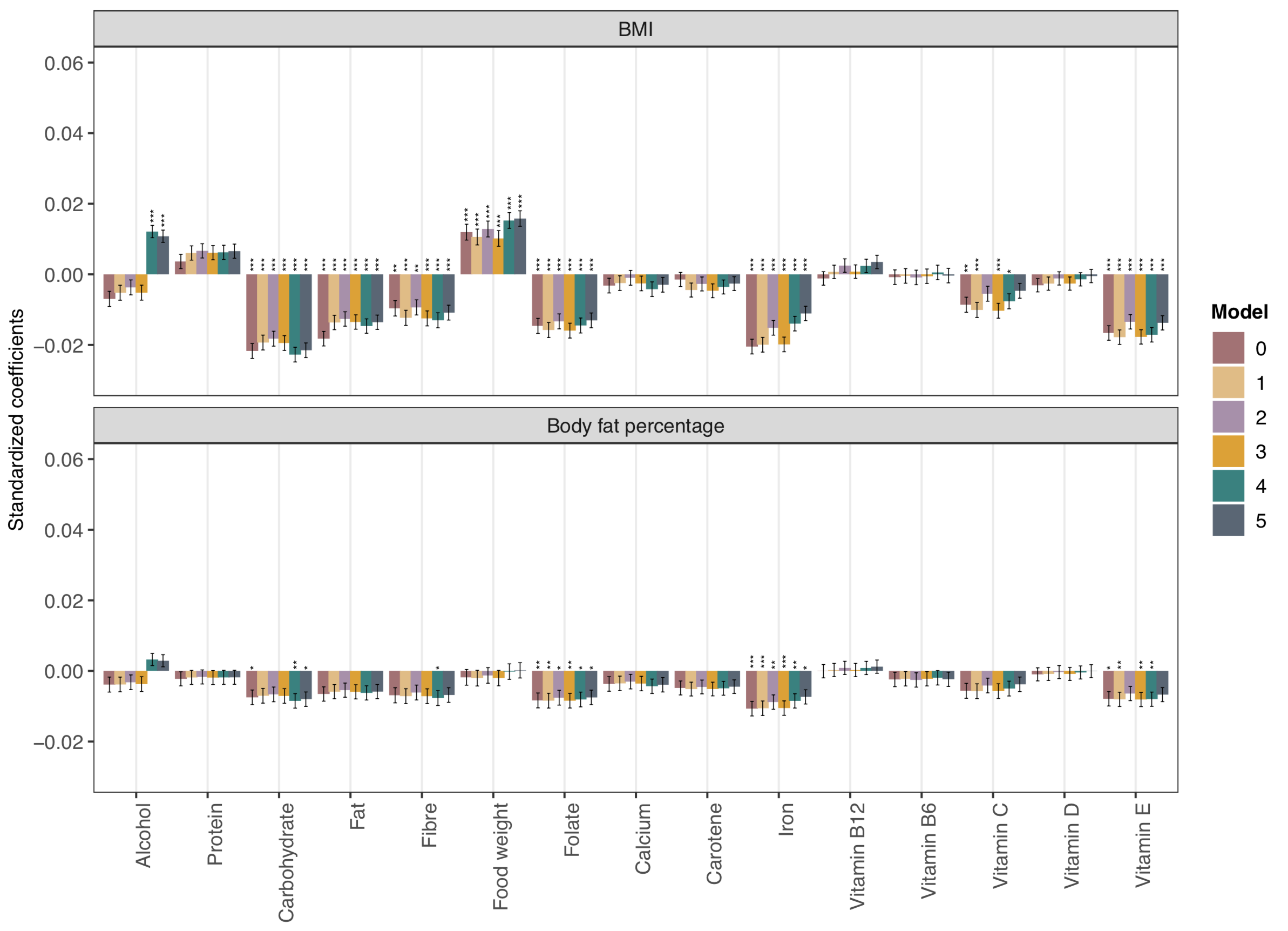

Results are shown from linear mixed-effects model analyses. Y axis shows the standardised β coefficient estimate (measured in units of standard deviation). PGSs and nutrient intakes were standardised with the figure showing the effect of a one standard deviation increase in PGS on nutrient intake. Colours represent the different models:

Model 0 - Sex, age and PC 1-6

Model 1: Sex, age, PC 1-6, special diet and typical diet yesterday

Model 2: Model 1 + socioeconomic status and educational attainment

Model 3: Model 1 + Physical activity

Model 4: Model 1 + diagnoses and medication that affect food intake, smoking and alcohol consumption

Model 5: All fixed effects

Error bars represent standard errors and asterisks indicate statistically significant estimates. Bonferroni-corrected p value thresholds: ∗ = p < 0.05/132, ∗∗ = p < 0.01/132, ∗∗∗ = p < 0.001/132
